# The impact of COVID-19 vaccines on patients with immune thrombocytopenic purpura: A protocol for a systematic review and meta-analysis

**DOI:** 10.1101/2023.12.09.23298879

**Authors:** Yangyang Li, Demin Kong, Yicheng Ding, Jinhuan Wang

## Abstract

**Introduction:** Immune thrombocytopenic purpura (ITP) is characterized by a decrease in platelet counts and can be triggered by various factors, such as viral infections and vaccinations. Concerns have emerged regarding potential links between the vaccines for COVID-19 and the worsening of ITP. This systematic review aims to comprehensively assess the impact of COVID-19 vaccines on patients with ITP, including associated risks and outcomes.

**Methods and Analysis:** A thorough search will be conducted across multiple electronic databases, including PubMed, Web of Science, EMBASE, Cochrane Library, CNKI, Wan Fang, VIP, and CBM, to identify pertinent studies. This study will encompass randomized controlled trials, cohort studies, case-control studies, and case series that assess the effects of COVID-19 vaccines on individuals with ITP. The primary outcome will center on alterations in platelet count, while secondary outcomes will encompass the occurrence of thromboembolic events, bleeding complications, recurrence rate of ITP, impact of ITP exacerbation, and adverse events. The data will be synthesized and subjected to meta-analysis using Review Manager Software (RevMan) V.5.3. Additionally, subgroup analyses will be conducted to investigate potential sources of heterogeneity.

**Ethics and dissemination:** Since this study involves the analysis of previously published data, ethical approval is not required. The findings will be disseminated through publication in a peer-reviewed journal and presentation at relevant scientific conferences.

**PROSPERO registration number** CRD42023471315.

**Strengths and limitations of this study:**

- Assessing the overall impact of COVID-19 vaccines on patients with ITP
- Incorporating a variety of study types to perform a thorough analysis
- Implementing a standardized methodology to evaluate and mitigate bias in the studies included
- Possible restrictions in data accessibility and variations in heterogeneity among the studies considered
- The effects of different types of COVID-19 vaccines on patients with ITP may differ, leading to potential disparities in the outcomes.

## INTRODUCTION

The autoimmune disorder known as immune thrombocytopenic purpura (ITP) is characterized by a decreased platelet count,^1, 2^ which poses a risk of bleeding and has a significant impact on patients’ quality of life.^3, 4^ This condition can be triggered by various inflammatory factors, such as infections or vaccinations,^5^ and has been further complicated by the global COVID-19 crisis.^6–8^ This has raised concerns about the potential adverse effects of COVID-19 vaccination in individuals with ITP.^9, 10^

Recently, there has been a growing focus on cases or worsening of ITP associated with COVID-19 vaccination.^11^ Multiple case reports have documented new occurrences or relapses of ITP in patients who have received COVID-19 vaccination, as well as after being infected with the virus.^12–15^ Although there is still ongoing debate about the causal relationship between COVID-19 vaccination and new cases of ITP, there have been reports of 16 ITP episodes following vaccination.^16–18^ Therefore, it is strongly recommended to closely monitor individuals after vaccination, considering the observed exacerbation of ITP in some patients. Research has shown that ITP patients experience a significant 6.3% decrease in platelet counts compared to healthy controls after receiving COVID-19 vaccination.^19^ While SARS-CoV-2 vaccines are generally regarded as safe for patients with preexisting ITP, they may contribute to worsening thrombocytopenia, particularly for patients who have undergone splenectomy or received multiple therapeutic interventions.^20^ Despite confirmed cases of ITP development following COVID-19 vaccination, the exact relationship between vaccination and primary or secondary ITP remains unclear.^21^

The evaluation of COVID-19 vaccines and their efficacy in diverse patient groups, particularly those with underlying conditions like immune thrombocytopenic purpura (ITP), has become imperative due to their extensive distribution. Given the potential influence of COVID-19 vaccines on immune response, it becomes crucial to evaluate their impact on individuals affected by ITP. Hence, the objective of this systematic review and meta-analysis is to provide a comprehensive insight into the risks and outcomes associated with COVID-19 vaccination in patients with ITP.

## METHODS

The research procedure adheres to the guidelines specified in the PRISMA-P (Preferred Reporting Items for Systematic Reviews and Meta-Analysis Protocol) statement.^22^

### Inclusion criteria

#### Types of studies

The assessment of the impact of COVID-19 vaccines on patients with immune thrombocytopenia (ITP) encompasses randomized controlled trials (RCTs), cohort studies, case-control studies, and case series.

#### Types of participants

The eligibility requirements comprise case studies and collections of cases analyzing the association between ITP and COVID-19 infection, as mentioned in the citations.^23–26^ To define ITP, the literature will be utilized, specifically excluding any other potential causes or types of thrombocytopenia. Any studies conducted in languages other than English or those addressing different causes or types of thrombocytopenia apart from ITP will not be included in the analysis.

#### Types of interventions Experimental interventions

We aim to incorporate all COVID-19 vaccination treatment interventions, encompassing diverse categories, dosages, regimens, and manufacturers. Moreover, we will take into account the administration of COVID-19 vaccination with varying intervals and sequences.

#### Comparator interventions

Our research will explore the effectiveness of various comparator interventions, including:

1. Individuals who have not been administered the COVID-19 vaccine.
2. Participants who have received a placebo or sham vaccination.
3. Subjects who have undergone other vaccinations or taken different medications.
4. Individuals who have received alternate types of COVID-19 vaccines.
5. Participants who have received varying dosages or treatment schedules for the COVID-19 vaccine.

Types of outcome measures

#### Primary outcome

The main focus of the study will be on the alteration in platelet count as the primary outcome.

#### Secondary outcome

Secondary outcomes that will be evaluated comprise the occurrence of thromboembolic events, complications associated with bleeding, recurrence rate of ITP, the impact of ITP exacerbation, and any adverse events.

### Search strategy

#### Electronic searches

In order to carry out an extensive investigation, relevant terms and keywords will be utilized to search significant electronic databases like PubMed, Web of Science, EMBASE, Cochrane Library, CNKI, Wan Fang, VIP, and CBM. Moreover, reference lists of pertinent studies and clinical trial registries will be thoroughly examined to identify potential eligible studies.Research papers published in both English and Chinese from the inception of the databases until October 6, 2024, were examined using the specified search terms: “idiopathic thrombocytopenic purpura”, “immune thrombocytopenic purpura”, “ITP”, or “immune thrombocytopenia”; “randomized controlled trial”, “controlled trial”, “random allocation”, “prospective study”, and “clinical trial”; as well as “COVID-19 Vaccines”, “Vaccines”, “Vaccination”, “Vaccin*”, and “sars-cov-2 vaccine”.

#### Searching other resources

For the purpose of discovering any additional pertinent research, we shall analyze the reference lists of the previously identified relevant systematic reviews, meta-analyses, and primary studies. Additionally, we shall establish correspondence with authorities and notable specialists active in the field of immunology and vaccine safety, in order to solicit information regarding any unpublished or ongoing trials. Furthermore, we shall conduct a meticulous exploration of the WHO International Clinical Trials Registry Platform (ICTRP), ClinicalTrials.gov, and other pertinent clinical trial registries, with a specific focus on ongoing or completed trials concerning the impact of COVID-19 vaccines on patients affected by immune thrombocytopenic purpura. In order to ensure a comprehensive review, we shall also undertake an exhaustive examination of grey literature using platforms like Google Scholar. This methodology endeavors to encompass all relevant studies within our review.

### Data collection and analysis

#### Selection of studies

The identified studies will be screened by two reviewers (YL and DK) independently. The screening process will be based on predetermined eligibility criteria. In case of any discrepancies, they will be resolved through discussion or consultation with a third reviewer (JW).

### Data extraction and management

To ensure the authenticity of the research, a standardized data extraction form will be utilized by two individual examiners (YL and DK) to adequately extract and arrange all pertinent data. In the event of any disparities, the original article will be referenced, and consultation with the principal investigator (JW) will be sought for resolution. The extracted data will encompass various components, including the primary author, publication year, journal source, geographical location, study methodology, sample size, patient age, ITP diagnostic criteria, intervention type, control group details, treatment frequency, outcome evaluation methods, identification of adverse effects, and any additional pertinent particulars.

### Assessment of risk of bias in included studies

The Cochrane Risk of Bias tool will be utilized by reviewers to evaluate 7 domains: random sequence generation, allocation concealment, blinding of participants and personnel, blinding of outcome assessment, assessment of incomplete data, selective outcome reporting, and identification of other sources of bias. The level of uncertainty, low risk, and high risk will be used to assess each domain of bias.

### Measures of treatment effect

The estimation of the impact for binary results shall be demonstrated as a risk ratio accompanied by a 95% CI. In the case of continuous outcomes, the estimation of the impact shall be presented as the mean difference accompanied by a 95% CI. If the identical outcome is assessed through various approaches, the magnitude of the intervention’s impact shall be communicated using the standardized mean difference accompanied by a 95% CI.

### Dealing with missing data

If possible, we will strive to establish contact with the authors of the study in order to obtain any missing data or seek clarification. To assess the potential impact of the missing data, the following strategies will be employed:

1. A worst-case scenario analysis will be conducted, wherein all participants with missing data will be considered as failures.
2. An extreme worst-case analysis will be performed, where participants with missing data in the experimental group will be deemed failures, while those with missing data in the control group will be regarded as successes.
3. An extreme best-case analysis will be undertaken, whereby participants with missing data in the experimental group will be considered successes, and those with missing data in the control group will be regarded as failures.

### Assessment of heterogeneity

We will examine the statistical heterogeneity by employing a χ^2^ test. Furthermore, we will measure the heterogeneity using the I^2^ statistic, which has a scale of 0% to 100%. Any p-value below 0.1 in the χ^2^ test or an I^2^ value exceeding 50% will indicate notable heterogeneity. To assess potential clinical heterogeneity, we will conduct prespecified subgroup analyses.

### Assessment of reporting biases

To evaluate seven domains, namely random sequence generation, allocation concealment, blinding of participants and personnel, blinding of outcome assessment, incomplete data assessment, selective outcome reporting, and other sources of bias, the Cochrane Risk of Bias tool shall be employed. Each domain shall undergo an assessment for levels of uncertainty, low risk, and high risk.

### Data synthesis

When examining studies that investigate the same treatment and produce similar outcomes in comparable populations, we will utilize meta-analysis to combine multiple trials and estimate the overall treatment effect. In order to pool continuous data, we will employ the inverse variance method, while for dichotomous data, we will use the Mantel-Haenszel method. In cases where there is low statistical heterogeneity, we will choose the fixed-effect model for data synthesis. However, if the p-value is below 0.1 or the I^2^ value exceeds 50%, we will employ the random-effect model to provide a more cautious estimation of the effect. All analyses will be conducted using Review Manager V.5.3 software. If it is not possible to conduct a meta-analysis, we will present a narrative summary of the individual study findings.

### Subgroup analysis and investigation of heterogeneity

To examine potential heterogeneity sources, such as variations in study designs, patient demographics, and COVID-19 vaccine types administered, subgroup analyses will be performed. These analyses will provide valuable insights into the potential impact of these factors on the outcomes of interest.

### Sensitivity analysis

To assess the robustness of the findings, sensitivity analyses will be carried out, which will involve evaluating the impact of individual studies on the overall results. These analyses aim to provide a deeper comprehension of the stability and reliability of the results obtained from the meta-analysis.

### Summary of findings table

To prepare the ’Summary of Findings’ table, we will utilize the Guidelines for Recommendation, Development, and Evaluation (GRADE)pro tool. This table will present an assessment score for the overall quality of evidence associated with each outcome.^27^ The evaluation of evidence quality will be conducted by two review authors independently, considering five grading criteria: study limitations, inconsistency, indirectness, imprecision, and publication bias. The assessment will assign one of four ratings: high, moderate, low, or very low. In case of any disagreements, consensus will be reached or a third reviewing author will be consulted for resolution.

### Amendments

We will provide the date of any amendment, a description of the change, and the rationale in the event of protocol amendments.

## ETHICS AND DISSEMINATION

This systematic literature analysis and meta-synthesis will uphold ethical principles and standards. Approval from the appropriate ethics review committee will be secured prior to conducting the investigation, ensuring the safeguarding and confidential handling of all individual data involved. Our aim is to disseminate our research findings comprehensively by publishing the final systematic literature analysis and meta-synthesis report upon the completion of the study. We plan to publish the outcomes in a peer-reviewed scientific journal and present them orally at academic gatherings to convey our research findings to the academic community and clinical practitioners. Additionally, we will disseminate the research results to the public through social media platforms and scientific communication channels in order to enhance awareness regarding the impact of COVID-19 vaccines on patients with ITP.

## Contributors

YL, DK and JW conceived the study; these three authors provided general guidance to the drafting of the protocol. YL and JW drafted the protocol. YD designed the search strategy. YL, JW, YD and DK drafted the manuscript. JW and YD reviewed and revised the manuscript. All authors have read and approved the final version of the manuscript.

## Patient consent

Not required.

## Provenance and peer review

Not commissioned; externally peer reviewed.

## Open Access

This is an Open Access article distributed in accordance with the terms of the Creative Commons Attribution (CC BY 4.0) license, which permits others to distribute, remix, adapt and build upon this work, for commercial use, provided the original work is properly cited.

See: http://creativecommons.org/licenses/by/4.0/

Article author(s) (or their employer(s) unless otherwise stated in the text of the article) 2018. All rights reserved. No commercial use is permitted unless otherwise expressly granted.

## Supporting information

Search strategy

## Data Availability

All the data generated in this study can be provided according to the reasonable requirements of the author. All the data generated in this study are included in the manuscript and are available online.

