## Supplementary material for "The impact of COVID-19 vaccines on patients with immune thrombocytopenic purpura: A protocol for a systematic review and meta-analysis": Search strategy

**MEDLINE：**

#1 Search related to "immune thrombocytopenic purpura":
(("Idiopathic Thrombocytopenic Purpura"[Mesh]) OR ("Thrombocytopenic Purpura, Idiopathic"[tiab]) OR ("Immune Thrombocytopenic Purpura"[Mesh]) OR ("ITP"[tiab]) OR ("Immune Thrombocytopenia"[tiab]))
#2 Search related to "COVID-19 Vaccines":
(("COVID-19 Vaccines"[Mesh]) OR ("Vaccines"[Mesh]) OR ("Vaccination"[Mesh]) OR ("Vaccin*"[tiab]) OR ("sars-cov-2 vaccine"[tiab]))
#3 Search related to "randomized controlled trial":
(("Randomized Controlled Trial"[Publication Type]) OR ("Controlled Clinical Trial"[Publication Type]) OR ("Random Allocation"[Mesh]) OR ("Prospective Studies"[Mesh]) OR ("Clinical Trial"[Publication Type]))
#4 #1 AND #2 AND #3

**PubMed:**

#1 Search related to "immune thrombocytopenic purpura":
(("Idiopathic Thrombocytopenic Purpura"[Mesh]) OR ("Thrombocytopenic Purpura, Idiopathic"[tiab]) OR ("Immune Thrombocytopenic Purpura"[Mesh]) OR ("ITP"[tiab]) OR ("Immune Thrombocytopenia"[tiab]))
#2 Search related to "COVID-19 Vaccines":
(("COVID-19 Vaccines"[Mesh]) OR ("Vaccines"[Mesh]) OR ("Vaccination"[Mesh]) OR ("Vaccin*"[tiab]) OR ("sars-cov-2 vaccine"[tiab]))
#3 Search related to "randomized controlled trial":
(("Randomized Controlled Trial"[Publication Type]) OR ("Controlled Clinical Trial"[Publication Type]) OR ("Random Allocation"[Mesh]) OR ("Prospective Studies"[Mesh]) OR ("Clinical Trial"[Publication Type]))
#4 #1 AND #2 AND #3

**EMBASE:**

#1 Search related to "immune thrombocytopenic purpura":
(‘idiopathic thrombocytopenic purpura’/exp OR ‘immune thrombocytopenic purpura’/exp OR ‘ITP’ OR ‘immune thrombocytopenia’)
#2 Search related to "COVID-19 Vaccines":
(‘COVID-19 Vaccines’/exp OR ‘Vaccines’/exp OR ‘Vaccination’/exp OR 'Vaccin* OR ‘sars-cov-2 vaccine’)
#3 Search related to "randomized controlled trial":
(‘randomized controlled trial’/exp OR ‘controlled clinical trial’/exp OR ‘random allocation’/exp OR ‘prospective study’/exp OR ‘clinical trial’/exp)
#4 #1 AND #2 AND #3

**Cochrane Library:**

#1 Search related to "immune thrombocytopenic purpura":
("idiopathic thrombocytopenic purpura" OR "immune thrombocytopenic purpura" OR

"ITP" OR "immune thrombocytopenia")
#2 Search related to "COVID-19 Vaccines":
("COVID-19 Vaccines" OR "Vaccines" OR "Vaccination" OR "Vaccin*" OR "sars-cov-2 vaccine")
#3 Search related to "randomized controlled trial":
("randomized controlled trial" OR "controlled trial" OR "random allocation" OR "prospective study" OR "clinical trial")
#4 #1 AND #2 AND #3

**VIP:**

（M=新型冠状病毒肺炎 OR COVID-19 OR 新冠肺炎 OR 冠状病毒病 OR 2019 冠状病毒病 OR 冠状病毒肺炎 OR 病毒性肺炎 OR 新冠 OR 新型冠状病毒 OR 2019-nCoV OR SARS-CoV-2 OR 2019 新型冠状病毒 OR 严重急性呼吸道综合征冠状病毒 2 型 OR 新冠病毒 OR 冠状病毒) AND (U=免疫性血小板减少症 OR 特发性血小板减少性紫癜) AND (R=随机 OR 随机对照 OR RCT)

**CNKI:**

TI=(新型冠状病毒肺炎+COVID-19+新冠肺炎+冠状病毒病+2019 冠状病毒病+冠状病毒肺炎+病毒性肺炎+新冠+新型冠状病毒+2019-nCoV+SARS-CoV-2+2019 新型冠状病毒+严重急性呼吸道综合征冠状病毒 2 型+新冠病毒+冠状病毒) AND SU=(免疫性血小板减少症+特发性血小板减少性紫癜) AND AB=(随机 OR 随机对照 OR RCT)

**CBM:**

("新型冠状病毒肺炎"[常用字段] OR "COVID-19"[常用字段] OR "新冠肺炎"[常用字段]) OR "冠状病毒病"[常用字段] OR "2019 冠状病毒病"[常用字段] OR "冠状病毒肺炎"[常用字段] OR "病毒性肺炎"[常用字段] OR "新冠"[常用字段] OR "新型冠状病毒"[常用字段] OR "2019-nCoV"[常用字段] OR "SARS-CoV-2"[常用字段] OR "2019 新型冠状病毒"[常用字段] OR "严重急性呼吸道综合征冠状病毒 2 型"[常用字段] "新冠病毒"[常用字段] OR "冠状病毒"[常用字段]) AND ("免疫性血小板减少症"[常用字段] OR "特发性血小板减少性紫癜"[常用字段]) AND "随机对照试验"[文献类型]

**Wan Fang:**

(题名或关键词=新型冠状病毒肺炎 OR COVID-19 OR 新冠肺炎 OR 冠状病毒病 OR 2019 冠状病毒病 OR 冠状病毒肺炎 OR 病毒性肺炎 OR 新冠 OR 新型冠状病毒 OR 2019-nCoV OR SARS-CoV-2 OR 2019 新型冠状病毒 OR 严重急性呼吸道综合征冠状病毒 2 型 OR 新冠病毒 OR 冠状病毒) AND (主题=免疫性血小板减少症 OR 特发性血小板减少性紫癜) AND (摘要=随机 OR 随机对照 OR RCT)
